# Transfer learning for mortality risk: A case study on the United Kingdom

**DOI:** 10.1101/2024.10.25.24316112

**Authors:** Asmik Nalmpatian, Christian Heumann, Levent Alkaya, William Jackson

## Abstract

This study introduces a transfer learning framework to address data scarcity in mortality risk prediction for the UK, where local mortality data is unavailable. By leveraging a pretrained model built from data across eight countries (excluding the UK) and incorporating synthetic data from the country most similar to the UK, our approach extends beyond national boundaries. This framework reduces reliance on local datasets while maintaining strong predictive performance. We evaluate the model using the Continuous Mortality Investigation (CMI) dataset and a drift model to address discrepancies arising from local demographic differences. Our research bridges machine learning and actuarial science, enhancing mortality risk prediction and pricing strategies, particularly in data-poor settings.

## Introduction

In life insurance, accurate mortality risk prediction is essential for pricing and managing risks. However, this process is often hindered by data scarcity, particularly in underrepresented demographic segments or smaller niches of the market. Mortality events are infrequent, meaning data accumulates slowly, making it difficult for insurers to build robust predictive models. This lack of data can lead to unreliable risk assessments and pricing strategies, ultimately affecting profitability and customer affordability.

Transfer learning offers a promising solution to these challenges by leveraging models trained on data-rich countries and adapting them to data-poor environments. This allows insurers to generate reliable mortality predictions even when local data is unavailable. Previous studies, such as those by [1] and [2], have laid the groundwork for transfer learning in mortality risk prediction, but have primarily focused on scenarios with small volumes of target data. Additionally, much of the research has relied on deep neural networks (DNNs), which, while powerful, can be computationally intensive and require extensive fine-tuning, especially for small datasets [3, 4].

In contrast, gradient boosting machines (GBMs) offer a more efficient and interpretable alternative for transfer learning, particularly in cases where no target data is available. Despite their potential, GBMs have received less attention in the context of mortality risk prediction. Inspired by the success of machine learning (ML) models in clinical research [5–7], this study introduces a GBM-based transfer learning framework for predicting mortality rates in the UK, where no local life insurance data is available. By incorporating synthetic data from countries most similar to the UK, this approach demonstrates high predictive accuracy while reducing dependence on local datasets. To further enhance the model, we introduce a drift model to evaluate and correct discrepancies arising from demographic differences between countries.

This research not only extends the boundaries of transfer learning in actuarial science but also has broader implications for improving mortality risk prediction and pricing strategies in data-poor markets. Our study is guided by three primary research questions:

i. ***How can we estimate mortality rates in a country with no internal life portfolio data?*** This involves implementing a ML-based transfer method, focusing on the UK, and constructing a country similarity index using external data to identify relevant source countries.
ii. ***How accurate is the model, and how can a drift model address discrepancies between predicted and expected mortality?*** The accuracy of the transfer learning method is assessed using various metrics, with a drift model employed to explore factors contributing to discrepancies between transferred mortality tables and expected outcomes from the CMI dataset.
iii. ***How can additional variables beyond age and gender improve mortality risk predictions*** We investigate how the inclusion of additional variables can enhance the baseline mortality predictions, providing an application case to demonstrate improvements..

## Database and methodology

### Data

In our study, we rely on the open source Human Mortality Database (HMD) as our primary external data source. HMD offers age and gender-specific mortality rates for the overall population across various countries. However, our primary focus is not on estimating the mortality of the overall population in the UK. Instead, our goal is to estimate the mortality rates within the company’s own life insurance portfolio in the UK. It’s important to note that there are often differences between overall mortality rates and those within a specific portfolio, particularly due to the underwriting process in life insurance. To address this limitation, we leverage data from eight countries and establish connections to capture this discrepancy between overall and portfolio mortality rates. To ensure that the analysis accurately reflects the mortality patterns across different countries and within the company’s life insurance portfolio, our approach involves three different populations, as illustrated in Fig 1: the overall population specific to each country, the global insured population of the company, and the insured population of the company within a particular country.

**Fig 1.**
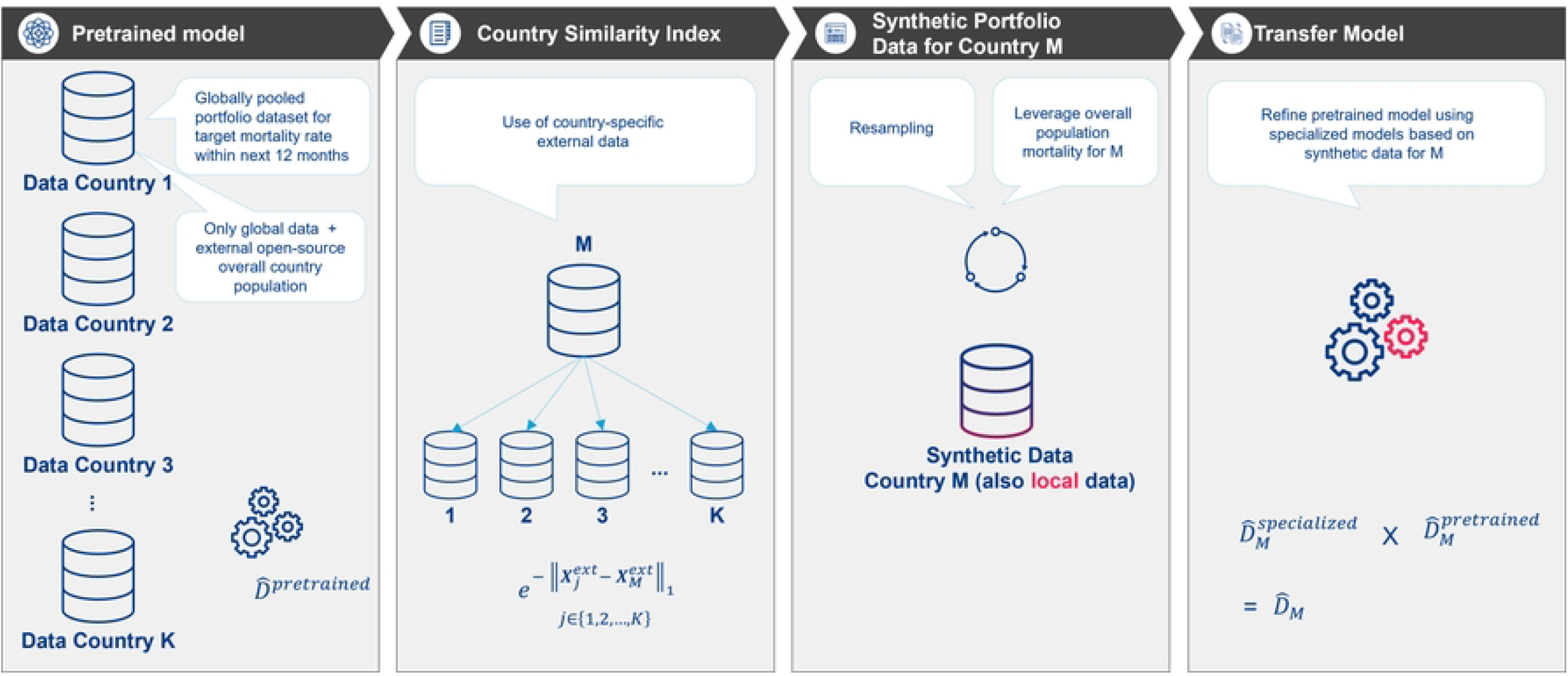
Illustration of targeted population segments across different datasets and models.

### Overall population

Age- and gender-specific overall population mortality rates from the HMD are retrieved for all countries. While these represent total population mortality, not insured population mortality, they bridge the gap between total and insured mortality, as it is the only feature we have available for the target country. To minimize yearly artifacts mortality rates from 2008 to 2018 were projected one year ahead using the Lee Carter model [8] (see Methodology section and S2 Appendix).

### Insured population

We utilize a pooled internal portfolio dataset from different countries to pretrain a GBM model [9] for predicting mortality rates for the insured population globally. This dataset incorporates common global characteristics shared across different countries, such as age, gender, sum assured, allowing for cross-country data comparison, and integrates the overall population mortality, yielding in a total of 9 global features. (see Methodology section and S1 Appendix).

The dataset includes policy data from a global primary insurer that was active during the specified period, totaling almost 10 million life-years of exposure and recording nearly 10,000 insurance claims (deaths). The data analysis was conducted in an aggregated form, grouped into distinct combinations of feature values, summarizing the deaths *D*_*j*_ and exposure *E*_*j*_ data for each unique combination features across all *j* = 1, …, *K* countries, in this case *K* = 8, the names of which have been withheld to maintain confidentiality. Four of the countries are located in Western Europe, three in Latin America, and one in Central and Eastern Europe.

Table 1 provides a detailed overview of *D*_*j*_, *E*_*j*_ and the total number of years *T*_*j*_ in country *j*, to give the main characteristics and distribution of the pooled dataset. This paper will analyse age and gender as internal features, while keeping other features used in the modeling anonymous for privacy reasons.

**Table 1.** Overview of death counts *D*_*j*_, exposure in life years *E*_*j*_, and total number of years *T*_*j*_ in country *j*.

| Country $j$ | $D_j$ | $E_j$ | $T_j$ |
| --- | --- | --- | --- |
| 1 | 1699 | 1 295 299 | 2013–2020 |
| 2 | 1291 | 1 686 299 | 2010–2020 |
| 3 | 494 | 815 795 | 2010–2020 |
| 4 | 1225 | 1 347 150 | 2017–2020 |
| 5 | 1816 | 1 825 901 | 2016–2020 |
| 6 | 2132 | 1 548 157 | 2016–2020 |
| 7 | 458 | 498 560 | 2017–2020 |
| 8 | 297 | 99 473 | 2015–2020 |
| Total | 9412 | 9 116 634 | 2010–2020 |

### Insured population in specific countries

In addition to the global features, including the overall population mortality of these countries, we include 12-16 local features from each country j, depending on local data availability, such as occupational class, which are not comparable across regions. After retraining the specialized GBM models on a total of 21-25 features, initialized by the pretrained model, we predict mortality rates for the portfolio of country M using a synthetic dataset tailored specifically for M. Our method for creating the synthetic dataset combines stochastic and rule-based techniques to bootstrap by resampling from the internal portfolio of K countries, while introducing variations to account for uncertainty [10] (see Methodology section and S1 Appendix).

#### Mortality of UK’s insurance population for evaluation

We utilize the ‘16’ series mortality tables from Working Paper 154 [11] for the evaluation purposes and the drift model, given the absence of an actual UK portfolio for comparison. These tables, derived from data from different UK life insurance companies, offer detailed insights into age, gender, smoking status, and curtate duration. To guarantee an impartial assessment and prevent undue complication, we consolidate the tables according to age and gender categories that correspond to population proportions.

#### External data for the Country Similarity Index

The Country Similarity Index seeks to measure the similarity between the target country *M* and the K (= 8) source countries in the internal dataset in terms of mortality and life insurance characteristics. We develop this by considering various indicators, selected based on prior research and expert input, adaptable to specific contexts. These indicators are categorized into three dimensions: Life Insurance Performance Indicators, Healthcare Statistics, and Overall Population Mortality. The details of these indicators are outlined in Table 2, with the methodology for their construction discussed in the subsequent subsection.

**Table 2.** Dimensions and items obtained from external sources for the construction of a Country Similarity Index related to mortality in life insurance.

| Item | Description and source |
| --- | --- |
| **1. Life Insurance Performance Indicators from OECD** |  |
| 1.1 Life Insurance Share | The ratio of gross life insurance premium to total gross premium, indicating the relative importance of life insurance compared to non-life insurance [12]. |
| 1.2 Density | The ratio of life insurance premiums to the whole population, measured in US Dollars [13]. |
| 1.3 Penetration | The level of development of the life insurance sector in a country, represented as a percentage [14]. |
| 1.4 Total Gross Premiums | Aggregate amount of premiums collected by life insurance companies in US Million Dollars [15]. |
| 1.5 Retention Ratio | Percentage of premiums retained by an insurance company rather than being transferred to reinsurers [16]. |
| **2. Healthcare Statistics** |  |
| 2.1 Health Care | Measured by The Health Index by Global Residence Index, providing an overall assessment of the healthcare system and general health of the local population. Ranges from 0 to 1, indicating low to high healthcare levels [17]. |
| 2.2 Retirement Pension | Country-specific Minimum Pensionable Age for Men obtained from Indicator Data [18]. |
| 2.3 Medical Staff per Capita | Number of physicians, nurses, and midwives per 1,000 people [19]. |
| 2.4 Hospital Beds per Capita | Number of hospital beds per 10,000 population [20]. |
| 2.5 Access to Basic Healthcare | Percentage of people with access to basic healthcare [21]. |
| 2.6 Healthcare Expenditure per Capita | Expenditure on healthcare per capita in US Dollars [22]. |
| 2.7 Risk of Impoverishing Expenditure for Surgical Care | Percentage of people at risk [23]. |
| **3. Overall Population Mortality** |  |
| 3.1 HMD Rates | Utilizing also here HMD's age- and gender-specific mortality rates by country [24]. |

### Methodology

*General Setup:* Consider a scenario where *K* source datasets with aggregated sample size *n*_*j*_ are collected from countries *j* = 1, …, *K* representing life insurance portfolios. The pooled dataset has total aggregated sample size 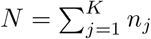. The objective is to estimate death counts *D* ∈ ℝ^*N*^ relative to exposure. The feature set *X* ∈ ℝ^*N ×p*^ comprises global features *X*^global^ that are comparable and available across countries including the overall population from HMD and local features *X*^local^ that are specific to each country. Our challenge arises in estimating mortality rates *D*_*M*_ due to the lack of internal data. However, we do have access to external data that provides information about mortality rates in different countries, including *M*. So, the scenario we are dealing with is comparing what we know from this external data along with some internal data we have (which is not specific to *M*) to try to estimate mortality rates specifically for country *M*. Fig 2 is a visual representation of the transfer learning framework: From the pretrained global model to the refined mortality rate predictions for the target county *M* based on a synthetic dataset.

**Fig 2.**
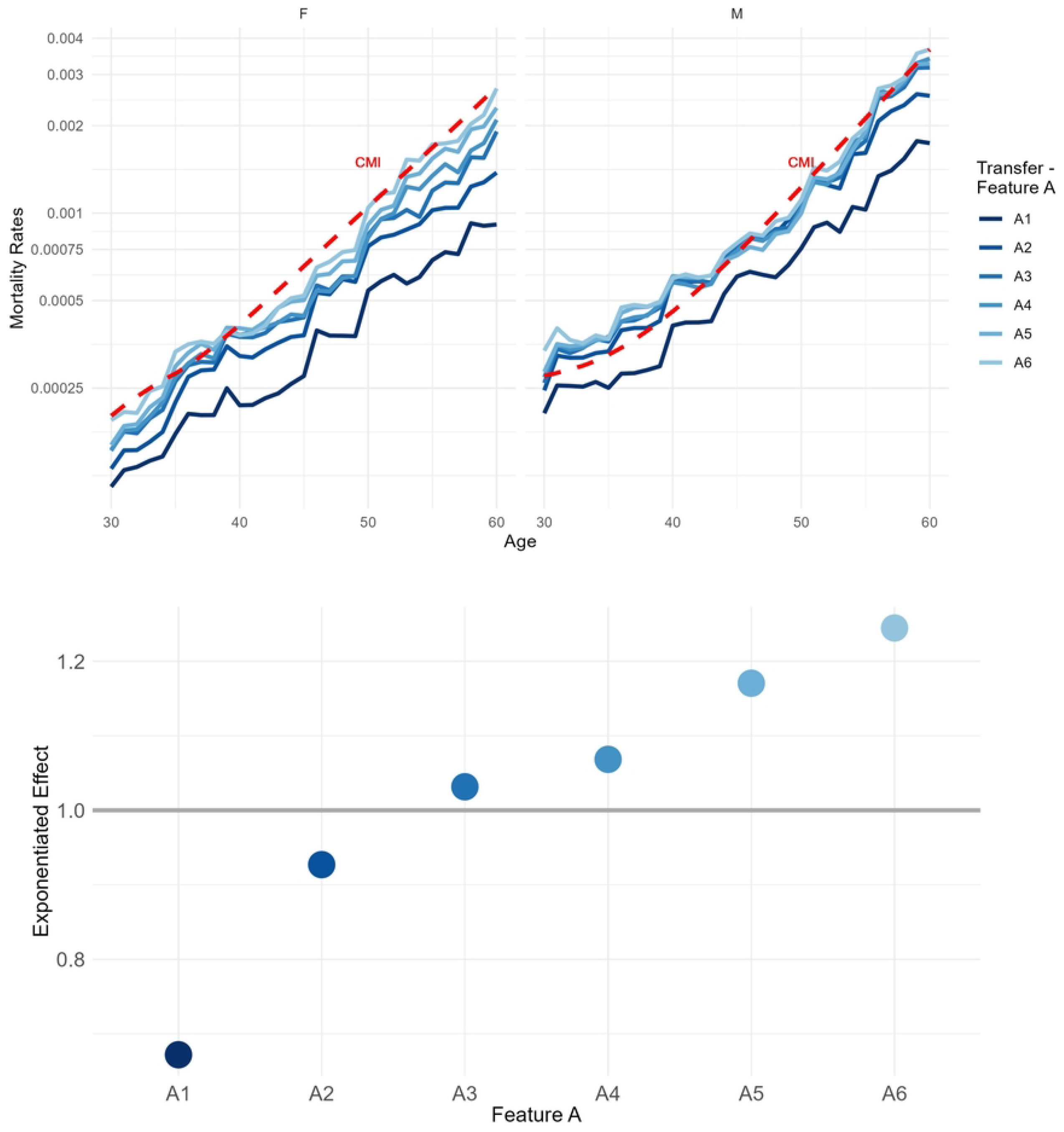
Framework sketch: Synthetic-data-based mortality predictions for target country M using a pretrained global mortality risk model.

#### Pretrained Model

Consider a broad category of risk prediction models, where the process of fitting the model involves using a loss function *L*(*γ*; *D, X*). With an estimated parameter vector 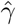 corresponding to the coefficients in a GBM, the predicted outcome is given by 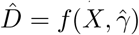. Specifically, we employ the negative Poisson log-likelihood function with Poisson distributional assumption. By minimising the expected loss function based on *X*^global^ we result in the parameter set estimate 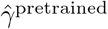 an thus predicted number of deaths 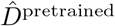. A detailed methodology for the GBM model is provided in S1 Appendix. Up to this point, a benchmark model has been developed without considering the country *M*. Previous work such as [25], [26], [4] and [27] characterize the similarity between the target model and the source models by a certain distance measure. Based upon this idea, we will generate a synthetic portfolio dataset *X*_*M*_ for country M, leveraging the similarity of the external data between the target population M and the source populations 1 to K (excluding M).

#### Country Similarity Index

To measure how similar the target country *M* is to the *K* source countries, we create a Country Similarity Index based on external insurance and mortality data *X*^ext^ ∈ R(*K*+1)*×Q*, with K number of source countries and 1 target country. In our application case, Q is equal to 13, larger than *K* + 1 = 9. These Q items, which are given in Table 2 apply to the entire population of a country, rather than internal data *X*, which specifically characterises the country’s insured population. After centering and scaling, the Manhattan distance between vectors 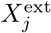 of each source *j* = 1, …, *K* and 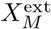 of target country *M* is calculated, as the sum of the absolute differences between corresponding components of vectors: 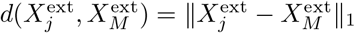. Finally, this results in a k-dimensional vector, representing the sum of itemwise distances between the *j* = 1, …, *K* and *M* across all Q items. The summation of distances over the countries is then transformed into the normed similarity score 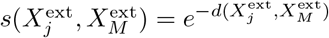 using the exponential function, so that the value range changes from [0, ∞) to (0, 1]. This transformation allows a similarity comparison rather than an absolute measure of distance, and becomes important later in the resampling stage to define the variance of the Gaussian distribution.

#### Synthetic Portfolio Data for Country M

In countries with no mortality data at all due to portfolio characteristics and size, synthetic data generation offers an efficient solution to address data limitations [29]. The process of producing mortality datasets that closely mimic actual data may comprise stochastic techniques [30], rule-based approaches set by human experts [31] or deep generative models (e.g., [32], [33]).

Assuming the known age and gender distribution for M, we resample feature combinations (rows) from the K datasets, encompassing both global and local features, along with the number of deaths, proportional to each similarity score 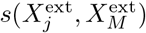 for *j* = 1, …, *K*. The overall population mortality of those countries has been substituted with the one of country M obtained from the HMD. To address potential unknown heterogeneity between j and M, we use a data augmentation technique with noise drawing inspiration from established practices (e.g., [34], [35]):

1. Metric Data: We introduce Gaussian noise with a mean *μ* of 0 and a standard deviation *σ* that is inversely proportional to the similarity score: 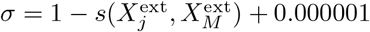. Higher similarity measure corresponds to a lower standard deviation, implying less noise is added to metric data.
2. Categorical Data: For categorical data, a noise level is drawn from 𝒩 (0, *σ*^2^), where again 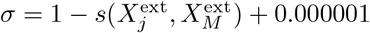. If the drawn value falls within a predefined interval around 0, the original value is retained, otherwise, a new value is drawn from the uniform distribution.

Finally, the synthetic dataset for the target country *M* is generated and contains the feature sets 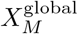 (including HMD) and 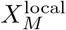 as well as the exposure *E*_*M*_ for country *M*. The estimation of death counts, denoted as 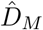, is required.

#### Transfer Model

Since the pretrained model excludes local factors like occupation class, which cannot be compared across countries, but may have significant impact on mortality, we calculate the specialized models with the local data on top. Each specialized model takes the output of the pretrained model from the first step and makes it more precise for that country. We find that incorporating local attributes during the latter phase of training offers optimal adaptability; this approach allows local nuances to be effectively integrated and, in cases where they are not applicable or transferable to the target country, they can be subsequently adjusted or mitigated. Initially, we utilize the global features of the synthetic dataset 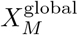 to generate preliminary predictions 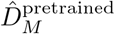 using a pretrained model. Subsequently, we enhance these predictions by employing the specialized GBM models tailored for countries *j* = 1, …, *K*. Through iterative boosting, the specialized model adjusts to the characteristics of the countries according to their similarity, thereby refining the mortality rate predictions. The final mortality rate predictions 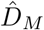 are determined by combining the specialized predictions 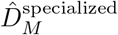 and the pretrained predictions 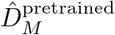 for all countries, as elaborated in the following Algorithm 1 and detailed out in S1 Appendix.

#### Agreement Metrics

Using several metrics we evaluate the agreement of transferred mortality rates 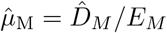 with the CMI mortality rates *μ*_cmi_, as proxy for expected UK mortality. Specifically, we employed Spearman correlation, cosine similarity and R-squared with centered expected versus predicted mortality rates. These metrics are defined as follows:

1. Spearman correlation:

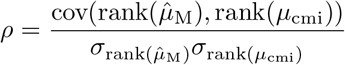
2. Cosine similarity:

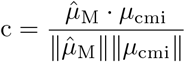
3. R-squared with centered actuals 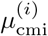 versus centered predicted vectors 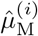:

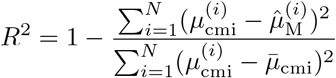

#### Drift Model Evaluation

We propose a drift model to evaluate the remaining disagreement by identifying and quantifying the drift drivers between target country’s expected mortality and the mortality rates transferred from other countries to M.

We assume a Poisson distribution for mortality counts in country *M*, denoted as *D*_*M*_ ∼ Poisson(*μ*_*M*_ · *E*_*M*_). Our analysis focuses on examining the discrepancy between the predicted mortality rate 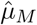 and the actual rate *μ*_cmi_ across various features or feature categories. This discrepancy, denoted as *δ*, serves as an indicator of the quality of transfer learning. We adopt the two-stage or residual model proposed by [36] to estimate *δ*:

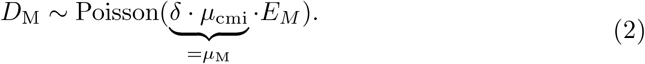

##### Algorithm 1

Algorithmic representation of the transfer framework

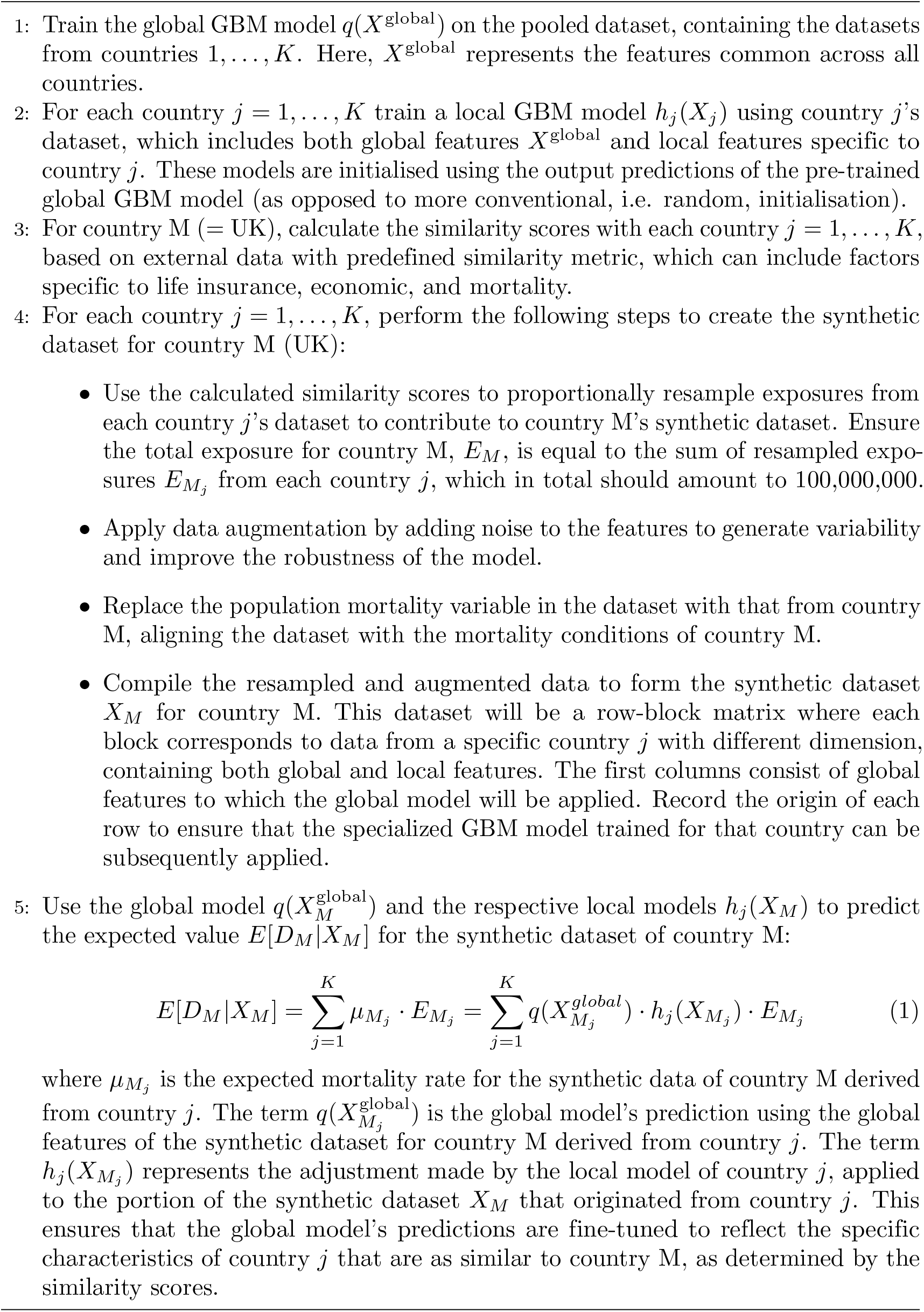

A Generalized Linear Model (GLM) is used with new exposure *D*_*cmi*_ = *μ*_cmi_ · *E*_*M*_, target 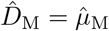 and model specification as follows [37]:

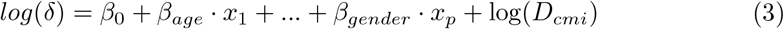

In the Poisson case, [38] demonstrated that the method is mathematically equivalent to using the ratio 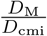 as target and *D*_cmi_ as weights:

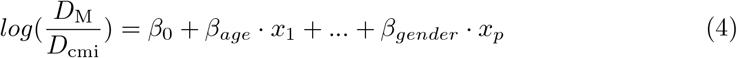

The validation of our approach, presented in Results section, includes comprehensive evaluation, such as its application to the UK insurance population and drift analysis from CMI mortality tables.

Due to exclusive usage of publicly available anonymized data (CMI and HMD) and aggregated, anonymized insurance data for model pretraining, there was no direct interaction with human participants, and no personally identifiable information was accessed. The insurance company data used for pretraining was provided in an aggregated and anonymized form, with no possibility of tracing back to any individual policyholder. No UK-specific data from the insurance company was used. The UK-specific results were derived entirely from publicly available data and a synthetic dataset generated for this study, with no real UK life insurance data being used.

Therefore, this study does not involve new data collection from human participants and participant consent was not applicable.

## Results

### Transfer learning application in the UK

The following section introduces the application of the transfer learning framework to the UK, where internal mortality data is unavailable. This analysis establishes the foundation for subsequent discussions and demonstrates a high level of agreement with expected outcomes.

The point of Fig 3 is to show the plausible transfer of knowledge from the countries to the UK, according to their similarity. It is clear that the degree of proximity is more pronounced in Europe, and therefore it makes more sense to resample from there than from the Latin American countries.

**Fig 3.**
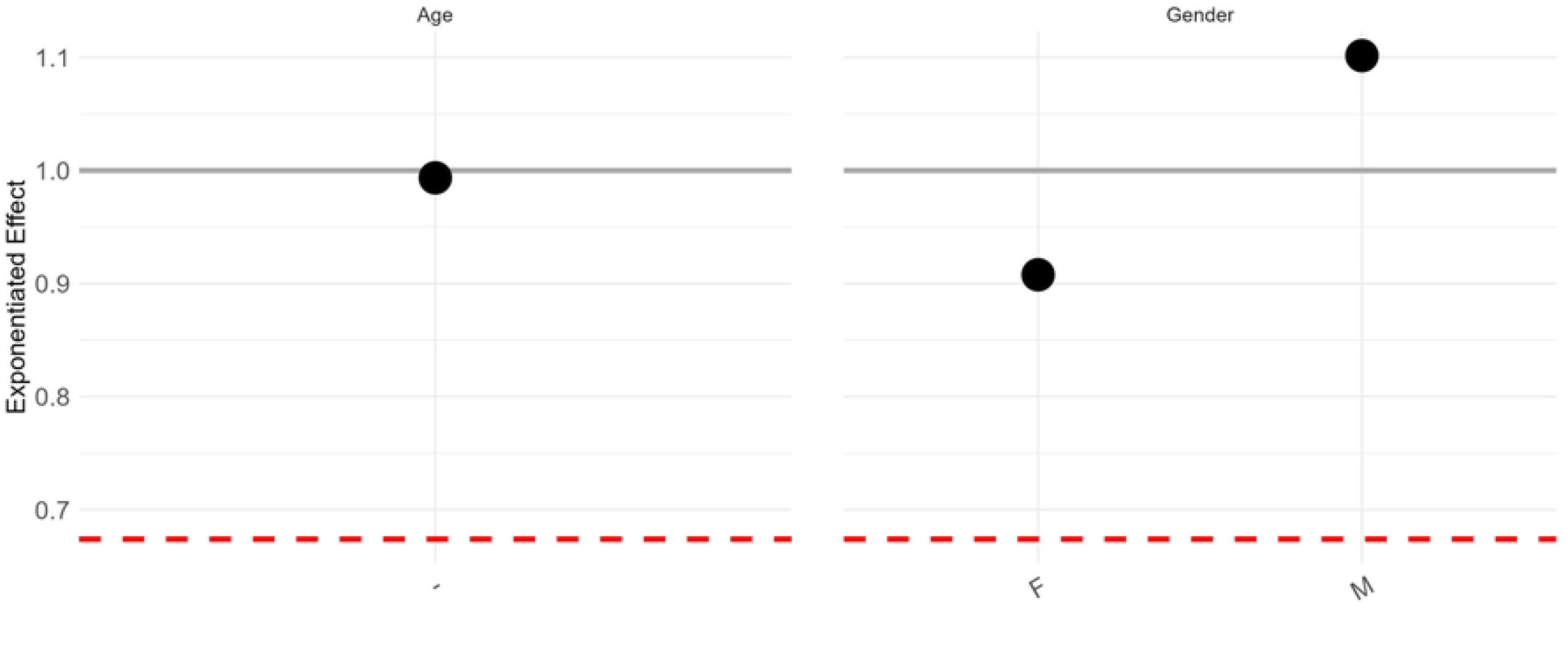
The composition of the exposure drawn from the countries for the synthetic dataset for the UK, proportional to the similarity score.

Fig 4 shows the predicted number of deaths for the UK based on the transfer model for age and gender. The remaining variables are not disclosed as they are considered to be insurer-specific and require confidential background information for proper interpretation. The categories with the highest exposure and claims are based on more original data, indicating greater reliability of the estimation and deserving of our focus.

**Fig 4.**
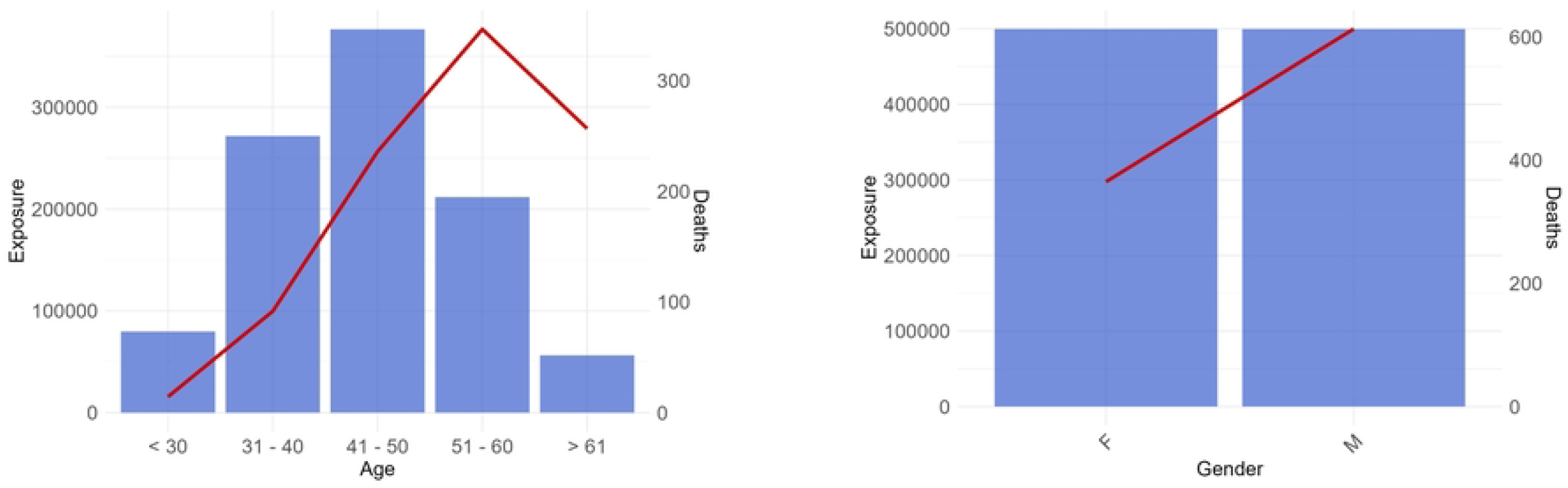
Exposure (bars) and predicted death counts (lines) by age and gender, derived from the synthetic-data-based transfer model. Age groups are defined retrospectively, and modeling is conducted using a metric scale. A. Age. B. Gender.

Furthermore addressing the second research question, we aim to evaluate the transfer model’s accuracy in matching the expected age-gender mortality rates using agreement metrics. CMI stands in for expected mortality rates in the UK’s insured population, given the lack of access to internal portfolio data. Despite differences in datasets and modeling, we regard CMI as a reliable proxy for UK policyholders’ actual mortality rates. The analysis focused on transferring insights about the relative mortality impact from different features in the data as opposed to producing an accurate estimate of the overall rate of mortality. This decision was made in part because it is expected that data will be available in the receiving country to estimate the overall rate of mortality, either from publicly available resources, or more likely from internal data that better reflects the specifics of the cohort being considered. Therefore, for evaluation purposes, we use Spearman correlation, cosine similarity, and R-squared as agreement metrics. These metrics do not consider the agreement of the difference in average mortality, ensuring objectivity in our evaluation.

Table 3 provides these measures not only for the UK but also for 8 other countries in the pooled dataset, as the transfer model’s predictive performance was also quantitatively examined for each of the 8 countries by pretraining the global model on the remaining seven. Given that the highest possible score is 1 for all metrics, we are within the highest acceptable range for the UK, as well as for the extended experiment.

**Table 3.** Evaluation metrics for different countries.

| Country | Spearman<br>Correlation | Cosine<br>Similarity | R-squared |
| --- | --- | --- | --- |
| UK | 0.9922 | 0.9878 | 0.9641 |
| 1 | 0.9221 | 0.9796 | 0.8641 |
| 2 | 0.8421 | 0.9253 | 0.8516 |
| 3 | 0.9711 | 0.9214 | 0.8912 |
| 4 | 0.9334 | 0.9658 | 0.8763 |
| 5 | 0.9242 | 0.9754 | 0.8773 |
| 6 | 0.8756 | 0.9564 | 0.8977 |
| 7 | 0.9360 | 0.8612 | 0.7822 |
| 8 | 0.9145 | 0.9700 | 0.8948 |

While the table indicates a high level of precision in estimating the age-gender mortality using the transfer learning framework, the following section proposes using the drift model to identify the cause of any remaining marginal discrepancies.

Fig 5 offers an initial insight into the disparities between the predictions of the transfer model and the CMI mortality rates, specifically examining age and gender. Despite an overall trend of underestimation in our estimates compared to CMI, our attention shifts to understanding the specific impacts of various features. Subsequently, we delve into the examination of age and gender as overlapping features present in both the predicted (transfer) and expected benchmark (CMI) mortality rates. To ensure monotonicity, it may be desirable to smooth the curves, i.e. to use them directly in pricing. We present our proposal for this in S3 Appendix, but in the main body we continue with the original version in order to remain faithful to the portfolio context and not to lose its specificity.

**Fig 5.**
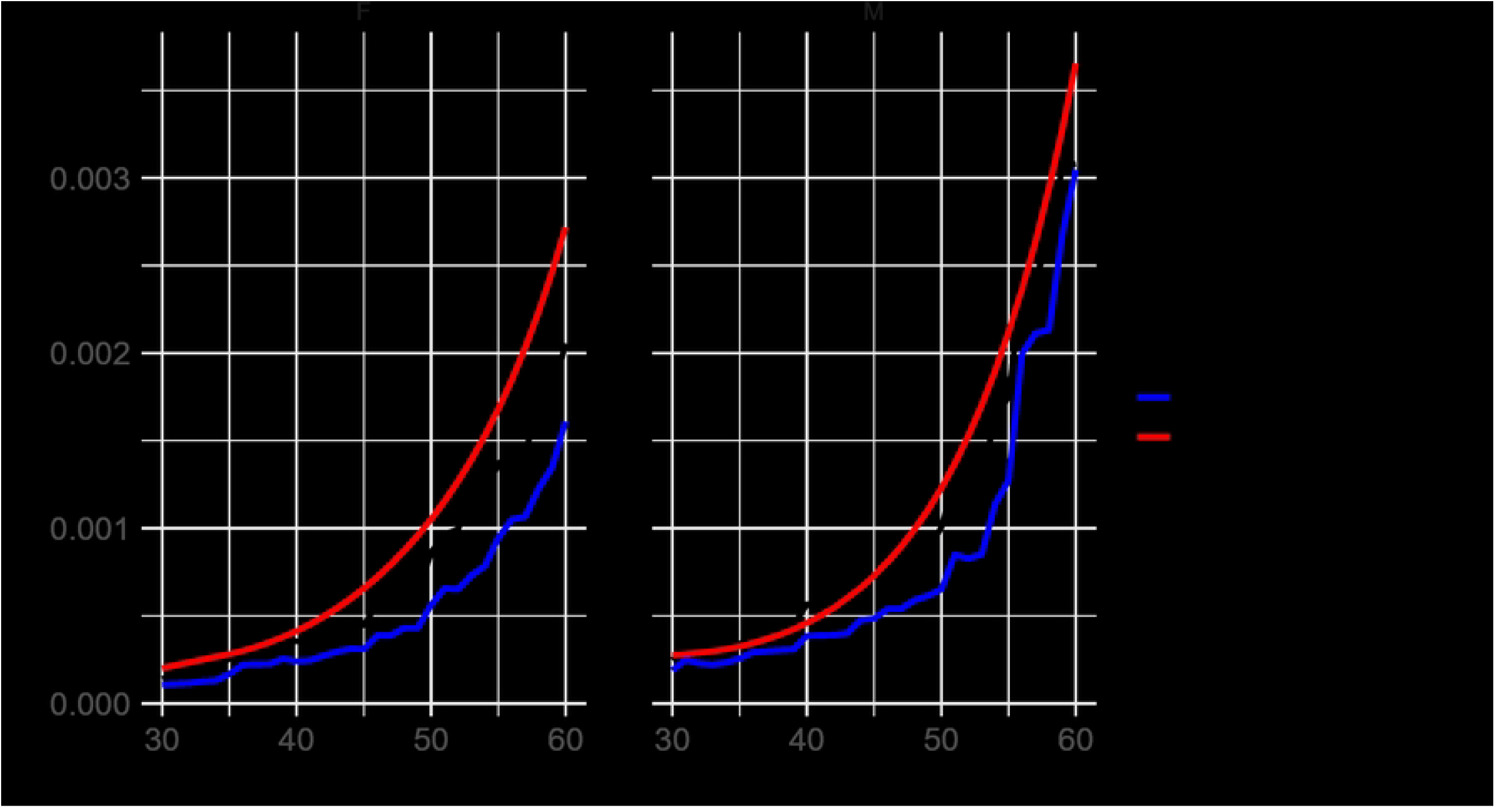
Comparison of UK mortality rates between Transfer Learning and CMI by age and gender. While transfer weighted by similarity score shows the above approach in black, the blue line shows the alternative of resampling only from the most similar country (MSC), which leads to a less accurate prediction.

Fig 6 illustrates the exponentiated coefficients of the drift model, offering insights into the relationship between the two mortality tables by quantifying deviations from the average ratio. The red dashed line at approximately 0.5 represents the exponentiated intercept 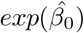, indicating the average ratio across all features. An exponentiated effect of 1 for a specific feature implies no impact on the ratio, suggesting effective capture of pattern differences between the source and target countries for that feature.

**Fig 6.**
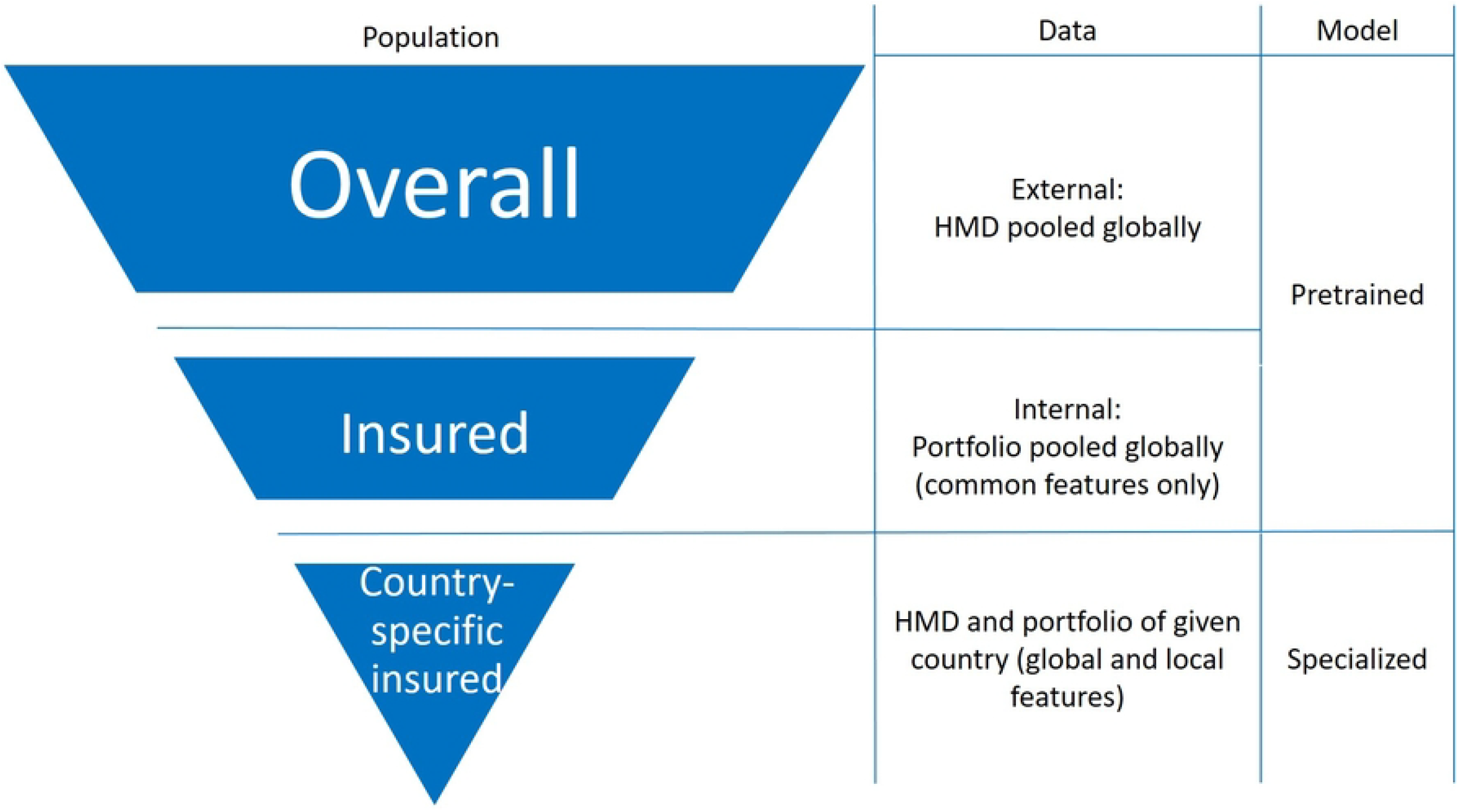
Exponentiated effects of age and gender on the ratio of transfer to CMI. The gray line represents the no-effect line, while the red dashed line is the exponentiated intercept.

The multiplicative effect of age in relation to the average ratio is approximately 1, indicating that age does not significantly influence the relationship between the transfer model and CMI. Although slight differences may exist in the age curve and average values, this suggests that the transfer learning framework effectively captured the shape variances between the other K (= 8) countries and the UK by age, resulting in a close replication of CMI. This successful matching of age curves is a critical finding for insurance purposes, and lends confidence to subsequent analyses. Despite being from a different country, the methodology achieves a close match to the expected age curve, providing a strong basis for further analysis.

Regarding gender-specific mortality risks, while both the transferred results and CMI indicate higher mortality rates for males than females, the transferred estimations may show slight discrepancies: males are slightly overestimated and females underestimated compared to the average mortality risk. However, these deviations appear minor and likely stem from cohort distinctions between CMI and internal data, as well as cultural differences between the primary reference countries and the UK’s insurance mortality data, possibly reflecting subtle cultural influences and evolving gender roles in different countries.

Building upon the strong alignment observed in the transfer learning process, the subsequent section investigates additional variables.

### Improving baseline mortality through additional variables in the transfer model

The drift model, which actually goes beyond age and gender, examines additional variables found in portfolio datasets but not included in the CMI. With the CMI serving as the insurer’s base table, the exponentiated effects estimated by the drift model for additional variables provide direct insight to insurers. This allows them to assess the potential impact of including these variables in the pricing model, and to determine possible loadings or discounts accordingly.

For example, considering Feature A with values A1, A2, A3, A4, A5, A6, absent from the CMI, Fig 7 (a) shows that the predicted mortality rates increase from A1 to A6. Consequently, the drift model’s exponentiated effects reveal that policies falling under A1 have a 33% lower mortality ratio compared to the average, while those under A6 exhibit a 24% higher ratio, both ceteris paribus. Therefore, a UK insurer may include an extra risk factor in their pricing strategy due to the relative risk of A1 being approximately 54% (67/124) of A6. This justifies a 33% loading for A6 policyholders. It is suggested that selection effects would significantly impact the risk profile. The estimation of all other variables is presented in S3 Appendix.

**Fig 7.**
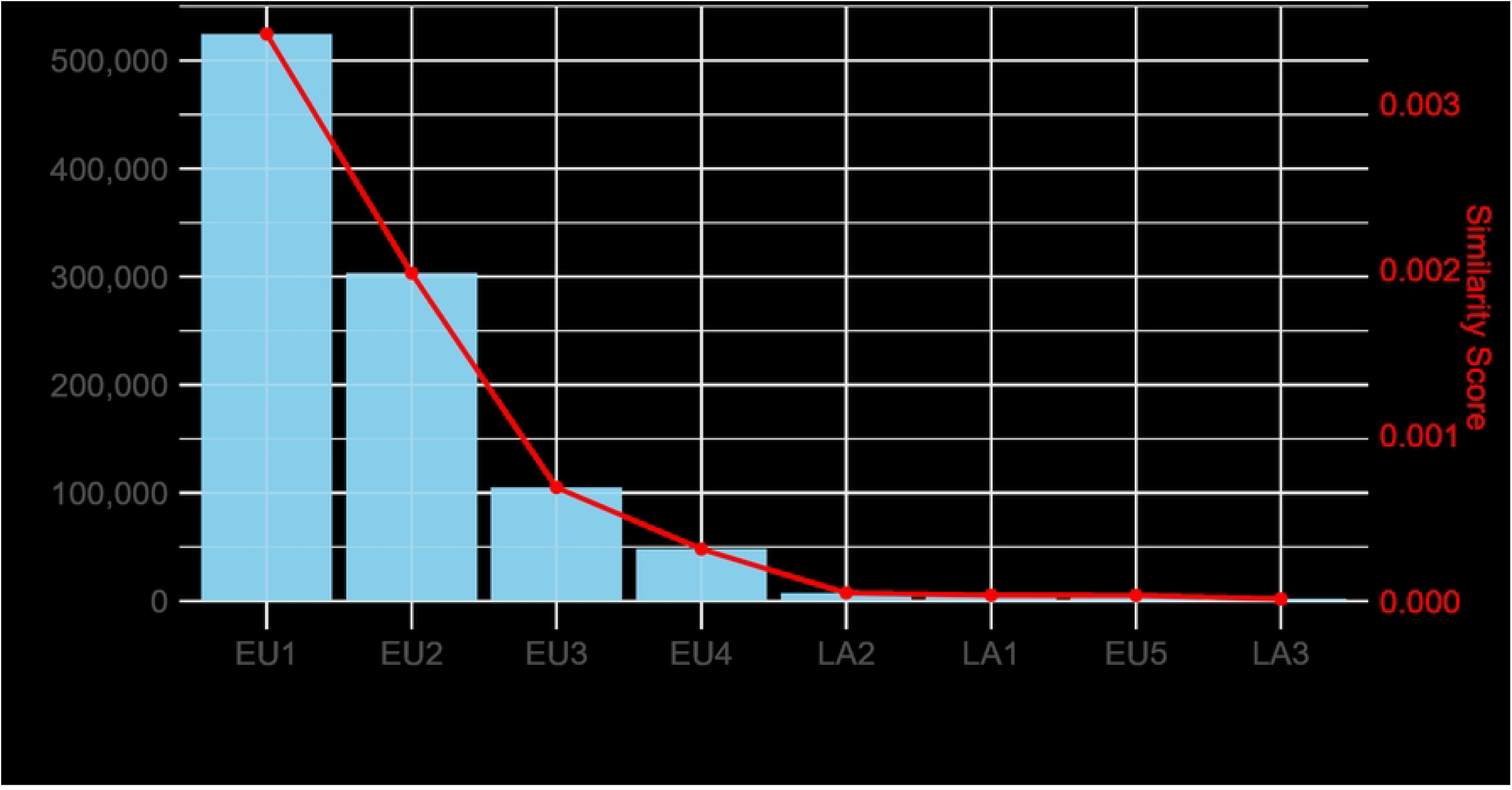
Feature A (with values A1-A6) evaluation as a risk factor for mortality. Transfer model results and evaluation of drift from CMI. A. The mortality rates for the UK are displayed on a logarithmic scale, segmented by Feature A. Red line represents CMI mortality rates. B. Exponentiated effects of Feature A on the ratio of transfer to CMI. The red line represents the exponentiated intercept, while the gray line represents the no-effect line.

In summary, the transfer learning framework effectively provides mortality risk predictions for the UK, leveraging a pretrained model from 8 other countries due to a lack of local mortality portfolio data, while refining the model using open-source UK total population mortality rates and data synthesized from the available countries accordingly to their similarity degree. While the model performs well with less culture-specific risk factors, discrepancies with CMI mortality tables highlight the need for evaluation using the drift model. This is essential for comprehensive risk assessment and to inform pricing strategies, particularly in scenarios where data is not available.

## Summary and outlook

This research presents a novel transfer learning framework designed to provide accurate mortality risk predictions for the UK, despite the complete absence of local mortality portfolio data. By leveraging pretrained and specialized models from eight other countries, along with UK population mortality rates obtained from open sources and synthesized data, we refine predictions for this data-scarce environment.

The framework establishes a solid foundation for mortality risk estimation and pricing, particularly benefiting small countries with insufficient data. Our predictive model shows strong agreement with the CMI mortality tables for age and gender, with only slight deviations detected via the drift model. Expert validation further supports the inclusion of additional variables to enhance mortality risk estimation.

The approach offers several practical benefits, including strong predictive performance, reduced reliance on local data, and lower computational demands, making it efficient for multi-centre studies. It simplifies the development and deployment of ML models by eliminating the need for extensive training data in each new country. Our findings suggest that transfer learning is particularly effective for factors that are less influenced by cultural differences, although it may experience drift when capturing local specificities.

While the reliance on synthetic data helps overcome data scarcity, it may introduce uncertainties, particularly when source countries differ demographically or economically from the target country. The effectiveness of the drift model also depends on the quality and similarity of external data used in the transfer learning process.

Future research could focus on addressing uncertainties in predictions by employing variance estimation to create confidence intervals. Incorporating additional socio-economic and regional factors may further improve mortality predictions. Expanding the framework to other regions and markets, especially those lacking sufficient local data, would provide valuable insights into its broader applicability. Testing the model in different settings could refine its use for life insurance product development in underserved demographic segments and emerging markets.

## Data Availability

The data are owned by a third party (insurance company) and authors do not have permission to share the data. The sources for external data is described.

## Supporting information

**S1 Appendix. Methodology details of pretraining and specialization**.

**S2 Appendix. Lee-Carter model**.

**S3 Appendix. Additional results of the drift model**.

## Notes

### Competing Interest Statement

The authors have declared no competing interest.

### Funding Statement

The author(s) received no specific funding for this work.

